# Benchmarking modular skills and fixed orchestration across six large language models in a meta-analysis task

**DOI:** 10.64898/2026.09.30.26363196

**Authors:** Yaqing Xiao, Yajun Gu, Pengfei Xia, Minying Deng, Shengyang Xie, Yingzhu Xing, Haoyue Cheng, Kun Zhu, Qianyu Yao, Fei Sun, Wei Chen, Yao Chen, Xueqian Wen, Yingyong Hou, Huimei Wang

**Author notes:** Equal first contribution (Yaqing Xiao, Yajun Gu, Pengfei Xia, Minying Deng). Equal second contribution (Shengyang Xie, Yingzhu Xing).

## Abstract

**Background:** Large language models (LLMs) are increasingly used to support systematic review and meta-analysis, but complex evidence flow, trial-family identification, study-level data extraction, and statistical execution remain vulnerable to omissions, untraceable data paths, and scientifically flawed steps. It remains unclear whether gains are driven by modular skills or by fixed orchestration, and whether these effects differ across base models.

**Methods:** We conducted a controlled, multi-model benchmark in which six LLMs completed the same randomized controlled trial (RCT) meta-analysis task under three conditions: (A) no meta-analysis-specific skills; (B) autonomous access to modular meta-analysis skills; and (C) the same skills executed under a Quality-First fixed directed acyclic graph (DAG). Each model was run three times per condition (54 runs total). The primary outcome was the scientific performance score (SPS, 0–100), rated by two meta-analysis experts; one expert was masked to explicit model and condition labels during SPS scoring, and final scores were the arithmetic mean of the two raters. Inter-rater reliability was high (SPS ICC(A,2) = 0.991).

**Results:** Mean SPS was 43.50 (condition A), 55.65 (B), and 54.44 (C). Relative to unassisted models, modular skills improved mean SPS by +12.15 points (unadjusted 95% CI 3.15–21.16; Holm-adjusted *P* = 0.033). The combined modular-skills-plus-fixed-DAG condition showed a positive but not statistically significant difference relative to unassisted models (*C − A* = +10.94; Holm-adjusted *P* = 0.073) and no incremental gain over skills alone (*C − B* = *−* 1.21; Holm-adjusted *P* = 0.746). No condition-by-model interaction was detected (*P* = 0.801).

**Conclusions:** In this benchmark task, access to modular skills improved mean scientific performance relative to unassisted execution; fixed orchestration of the same skills did not provide a detectable incremental benefit. The findings are task-specific and do not establish general effects for other meta-analysis settings, models, or orchestration designs.

## 1 Introduction

Systematic review and meta-analysis comprise interdependent steps, including screening, trial-family de-duplication, outcome definition, study-level data extraction, risk-of-bias assessment, statistical input, and visualization of pooled results. These steps are tightly coupled: a single upstream error in screening or extraction can propagate into the pooled estimate and the final conclusion [1, 2]. LLMs possess strong text comprehension and code-generation capabilities, and prior work has shown that foundation models can encode clinically relevant knowledge and assist with biomedical workflows [3, 4]. However, native LLM outputs remain vulnerable to omissions, untraceable data, and statistically flawed analysis paths, and open-ended execution does not guarantee that reported results are reproducible or scientifically correct [5, 6].

Two scaffolding strategies have emerged for improving reliability [8–11]. Modular skills decompose a complex task into reusable functional units that a model can select and connect according to task needs; recent skill-based agent research has shifted attention toward reusable skills, skill routing, dependency-aware retrieval, and workflow-level benchmarking [12–15]. An orchestrator, in contrast, prescribes execution order, dependencies, and conditional nodes through a fixed directed acyclic graph (DAG), in principle reducing omissions and broken data chains. The two strategies carry different costs: free orchestration relies on the model’s own planning ability, whereas fixed orchestration may introduce unnecessary nodes, increase context load, and propagate errors when early decisions are wrong.

Prior evaluations have frequently varied tools, prompts, workflows, and base models simultaneously, making the source of any performance gain difficult to attribute [16]. Single-model or single-run evaluations, particularly those relying only on automatic metrics, cannot capture model-to-model heterogeneity or run-to-run stability. For meta-analysis tasks in particular, fluent output is not equivalent to scientific correctness: claims must be checked against study-level gold standards and expert judgment rather than language-quality proxies [16]. We therefore varied skill availability and orchestration independently while holding the task, inputs, and evaluation constant. We evaluated six LLMs under three controlled conditions that isolated baseline model performance, access to modular meta-analysis skills, and fixed orchestration of the same skills. We hypothesized that modular skills would improve scientific performance relative to unassisted models, while the incremental benefit of fixed orchestration and its dependence on the underlying model were evaluated separately. All runs were scored by two meta-analysis experts, with one expert masked to model and condition labels during SPS assessment.

## 2 Methods

### 2.1 Study design and overview

This was a controlled, multi-model, three-condition, repeated-run benchmark evaluation. The unit of analysis was the individual LLM run (*n* = 54), not the 20 clinical studies underlying the benchmark input. Six LLMs completed the same RCT meta-analysis task under three experimental conditions (A, B, C), with three independent runs per model per condition. The primary outcome was the scientific performance score (SPS). All runs were scored by two meta-analysis experts, one of whom was masked to explicit model and condition labels during SPS scoring; final scores were the arithmetic mean of the two experts. Figure 1 shows the Quality-First fixed orchestration workflow; the experimental design is described in the text. The study was not formally preregistered.

**Figure 1:**
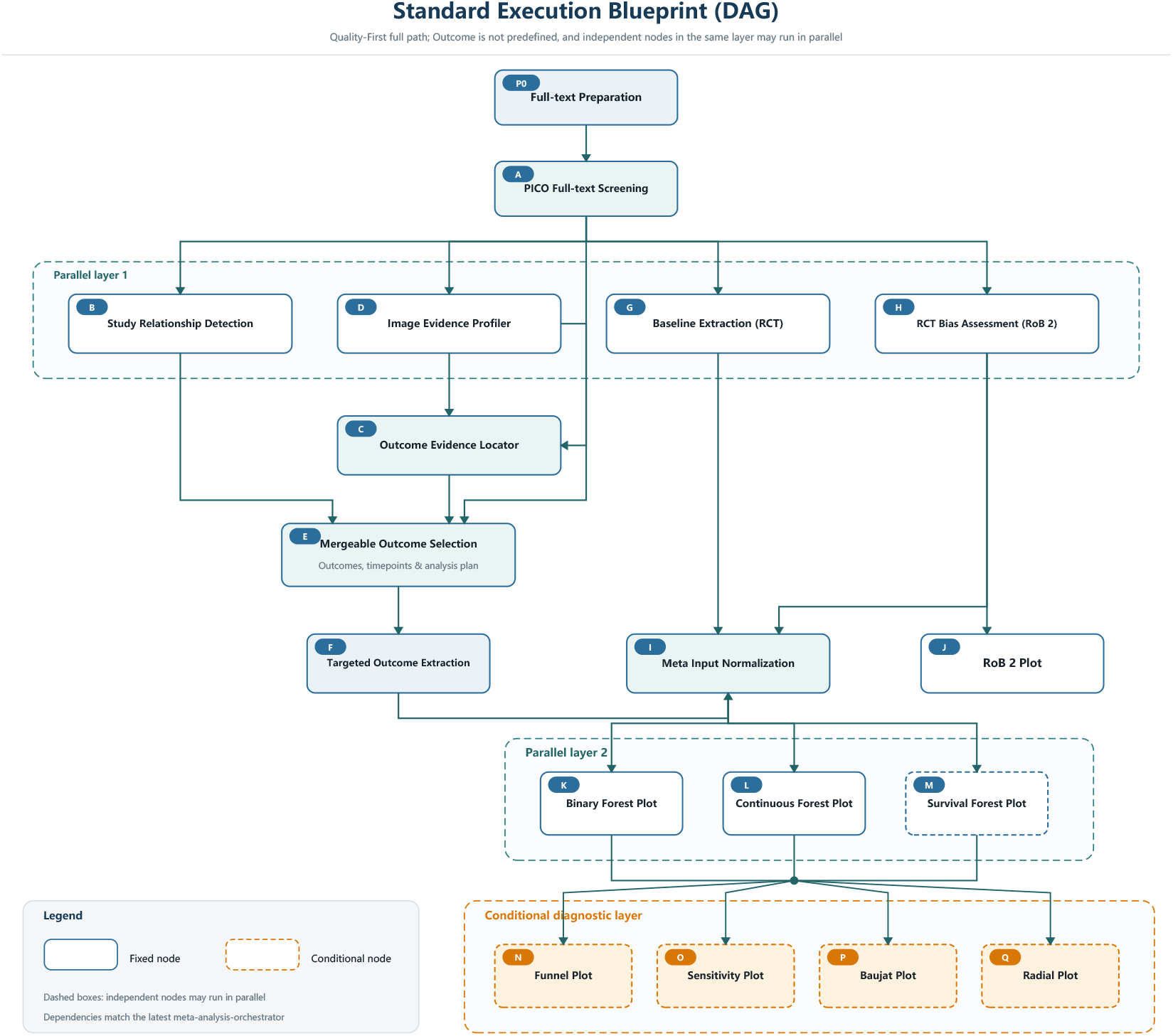
Fixed Quality-First orchestration workflow. The figure shows the 18-node skill workflow (P0, A–Q) spanning full-text preparation, re-screening, outcome selection, extraction, risk-of-bias assessment, standardization, meta-analysis, and diagnostic plots, with parallel layers, serial dependencies, and conditional nodes.

### 2.2 Benchmark meta-analysis task

The benchmark task was a pre-specified RCT meta-analysis comparing robotic-assisted with conventional laparoscopic surgery for colorectal cancer. The population comprised patients with colorectal cancer. The intervention was robotic-assisted surgery, the comparator was conventional laparoscopic surgery, and eligible study designs were limited to randomized controlled trials [2]. The input comprised 20 fully parsed study packages derived from the full texts of published RCT reports; the packages were reused from an existing parsed corpus, and PDFs were not re-downloaded or re-parsed for this study. The task specified the study design, intervention, comparator, and four task outcomes. Three outcomes served as formal SPS endpoints: conversion to open surgery, operative time, and number of lymph nodes harvested. Postoperative complications were collected but evaluated only descriptively because they were not pre-specified by type, severity, or follow-up window in the original task, and they did not contribute to SPS scoring, score caps, or scientific-safety classification.

### 2.3 Experimental conditions

The three experimental conditions differed only in the availability of meta-analysis-specific skills and in orchestration. In condition A, models completed the task without access to meta-analysis-specific skills. In condition B, models could autonomously select, order, and connect individual modular skills. In condition C, models had access to the same skills as in condition B but were required to execute them under the Quality-First fixed DAG. All other inputs, permissions, and output requirements were identical across conditions. In the figures, these three conditions are labeled Groups A, B, and C.

### 2.4 Modular skills and the Quality-First DAG

The skill package covered the functional domains of the meta-analysis workflow: screening, trial-family identification, outcome location and selection, study-level extraction, risk-of-bias (RoB 2) assessment [17], data standardization, meta-analysis, and diagnostic plots. The Quality-First DAG defined parallel layers, serial dependencies, and conditional nodes that governed when downstream steps could be executed. Detailed definitions of the 18 nodes (P0 and A–Q), their input/output schemas, and the complete DAG are provided in the Supplementary Methods (Table S1); the full workflow is shown in Supplementary Figure S1 and the skill data flow in Supplementary Figure S2.

### 2.5 Models, execution environment, and repetitions

Six LLMs were included: GPT-5.5, Claude Sonnet 4.5, GLM-5.2, DeepSeek-V4-Pro, Kimi-K2.6, and MiniMax-M3. Models were selected pragmatically to cover widely used systems available to the study team across multiple providers and deployment ecosystems; they were not intended to represent a systematic sample of all available LLMs. Each model ran the task three times under each condition (nine runs per model; 54 runs total). Run labels rep01–rep03 were used as the pairing basis in the statistical analysis: runs were paired by model and replicate number, yielding 18 paired observations per comparison. Context-window sizes were 1M tokens for GPT-5.5, GLM-5.2, DeepSeek-V4-Pro, and MiniMax-M3 and 256K tokens for Claude Sonnet 4.5 and Kimi-K2.6. With the exception of Kimi-K2.6 (temperature = 1.0, top_p = 0.95), all models used temperature = 0.2 and top_p = 1.0. Models were executed on the OpenCode 1.18.9 platform with Python 3.12 and R 4.4.0 (Table 1). Up to three retries were permitted on failure. Models were executed in parallel, and there was no information exchange between model runs.

**Table 1:** Models and controlled execution environment. All models were executed in parallel with independent contexts on the OpenCode 1.18.9 platform with Python 3.12 and R 4.4.0; up to three retries were permitted on failure.

| Model | API Provider | Context | Temperature | Top_p | Retry Limit | OpenCode | Python | R |
| --- | --- | --- | --- | --- | --- | --- | --- | --- |
| GPT-5.5 | OpenAI | 1M | 0.2 | 1.0 | 3 | 1.18.9 | 3.12 | 4.4.0 |
| Claude Sonnet 4.5 | Anthropic | 256K | 0.2 | 1.0 | 3 | 1.18.9 | 3.12 | 4.4.0 |
| GLM-5.2 | Zhipu AI | 1M | 0.2 | 1.0 | 3 | 1.18.9 | 3.12 | 4.4.0 |
| DeepSeek-V4-Pro | DeepSeek | 1M | 0.2 | 1.0 | 3 | 1.18.9 | 3.12 | 4.4.0 |
| Kimi-K2.6 | Moonshot AI | 256K | 1.0 | 0.95 | 3 | 1.18.9 | 3.12 | 4.4.0 |
| MiniMax-M3 | MiniMax | 1M | 0.2 | 1.0 | 3 | 1.18.9 | 3.12 | 4.4.0 |

### 2.6 Gold standard and SPS

The SPS comprised five weighted dimensions summing to 100 points: full-text re-screening and analysis eligibility (15 points), study-level data and analysis paths for the three formal outcomes (30 points), meta-analytic methods and results (30 points), core artifacts and analysis completeness (15 points), and verifiability and reporting completeness (10 points). A study-level gold standard was constructed that allowed multiple defensible branches, for example the inclusion or exclusion of non-Chinese/non-English studies according to a pre-specified language policy, the inclusion of continuous data with or without transformation, and the mutually exclusive handling of REAL/Tang criteria or parent-study/sub-study reports. For each acceptable branch, the corresponding study set, meta-analysis input, pooled effect, and tolerance ranges were predefined, and extraction was scored against study-level reference data with these tolerances. Scientific safety/risk levels were assigned by both experts according to explicit criteria based on the scientific-safety bottom line, as detailed in the Supplementary Methods (Section S8). The gold standard was established and reviewed by the same expert (Expert 1), a long-term meta-analysis researcher. The complete rubric (Table S2), reference data, scenarios, tolerances, and the definitions of the defensible gold-standard branches are provided in the Supplementary Materials (Section S5).

### 2.7 PFS and exploratory outcomes

Group-specific Protocol-following scores (PFS-A, PFS-B, PFS-C), each scored out of 15 points, measured how completely each run followed the process requirements of its own condition. PFS-A assessed compliance with the requirement not to use meta-analysis-specific skills, task boundaries, and method instructions; PFS-B assessed the actual invocation, outputs, and upstream/downstream transfer of individual skills; and PFS-C assessed the Quality-First orchestrator, DAG dependencies, and data transfer at key nodes. These three scores are condition-specific: they are not directly comparable across conditions and are not summed with the SPS. Postoperative complication handling was assessed descriptively in a post hoc analysis of whether the model identified inconsistencies in outcome definitions or time windows and whether it adopted stratification, unified definitions, stopped pooling, or flagged human review. This exploratory assessment was not compared with reference complication data and did not contribute to SPS or PFS; no hypothesis testing was applied.

### 2.8 Expert rating, masking, and adjudication

All 54 runs were scored independently by two experts. Expert 1 was aware of models and conditions and participated in gold-standard construction. Expert 2 was masked during SPS scoring, and runs were presented under random identifiers B001–B054. Explicit model and condition labels were removed from run texts and text embedded in PNG files; time metadata and file timestamps were also removed. The identifier mapping was generated from a confidential seed and path-based SHA-256 ordering, and runs were delivered in nine masked rating batches. Source and masked directories were cross-checked by SHA-256, with de-identification logs retained. PFS scoring was not masked because process evidence (skills and DAG logs) had to be visible to the rater. The design is therefore accurately described as one masked and one unmasked expert, rather than double-blinding. Final continuous scores were the arithmetic mean of the two experts. Risk-level classifications were recorded for both experts and summarized by raw agreement and weighted *κ* (Section 3.2). The mean expert difference (Expert 1 minus Expert 2) was computed to accompany the ICC.

### 2.9 Statistical analysis

The unit of analysis was the individual run. Conditions were summarized as mean *±* between-run SD. The primary comparison was *B − A*; key secondary comparisons were *C − A* and *C − B*. Comparisons were evaluated with two-sided paired *t* tests, with pairs defined by model and replicate number, and *P* values were adjusted across the three comparisons using Holm’s procedure. Reported 95% confidence intervals were not adjusted for multiple comparisons. A two-factor model, SPS *∼* condition *×* model, estimated the condition and model main effects and their interaction; model-specific contrasts were described only in the context of the interaction. Rater reliability was assessed with two-way mixed, absolute-agreement intraclass correlation coefficients (ICC(A,1) and ICC(A,2)) with 10,000 bootstrap 95% CIs [18, 19], together with the mean expert difference; risk-level agreement was summarized by raw agreement and weighted *κ* [20]. Exploratory analyses included within-condition PFS–SPS correlations (no pooled slope), descriptive run-level direction counts, and descriptive complication handling; all exploratory results are labeled as such. Analyses were performed with R 4.4.0.

## 3 Results

### 3.1 Run completion and rating dataset

All 54 runs produced evaluable output: 18 per condition and 3 per model per condition. Both experts completed SPS and PFS scoring for all 54 runs.

### 3.2 Expert agreement supports the stability of SPS measurement

SPS inter-rater reliability was high: ICC(A,2) = 0.991 (95% CI 0.988–0.993) and ICC(A,1) = 0.982. PFS reliability was similarly high (ICC(A,2) = 0.988; ICC(A,1) = 0.977). Expert 1 rated on average 1.99 points higher than Expert 2 (95% CI 1.23–2.75), indicating a systematic rater-severity difference despite high absolute agreement; scoring differences did not show obvious condition or model dependence (condition *P* = 0.342; model *P* = 0.413), and no batch drift was detected across the nine masked-rating batches (Spearman *r* = 0.093, *P* = 0.509; batch effect *P* = 0.525). The raters agreed on risk level in 81.5% of runs (44/54), with a weighted *κ* of 0.867; in all 10 discordant cases, Expert 2 assigned a higher risk level (8 low-to-moderate, 2 moderate-to-high).

### 3.3 Modular skills improve SPS relative to unassisted models

Mean SPS was 43.50 (SD 15.37) in condition A, 55.65 (SD 17.35) in condition B, and 54.44 (SD 19.27) in condition C. The primary comparison *B − A* was +12.15 (unadjusted 95% CI 3.15–21.16; Holm-adjusted *P* = 0.033), a statistically significant improvement with modular skills relative to unassisted models. *C − A* was +10.94 (unadjusted 95% CI 0.77–21.12; Holm-adjusted *P* = 0.073), a positive but not statistically significant difference after adjustment. *C − B* was *−*1.21 (unadjusted 95% CI *−* 8.94 to 6.52; Holm-adjusted *P* = 0.746), providing no evidence of an incremental gain from fixed orchestration beyond skills (Figure 2).

**Figure 2:**
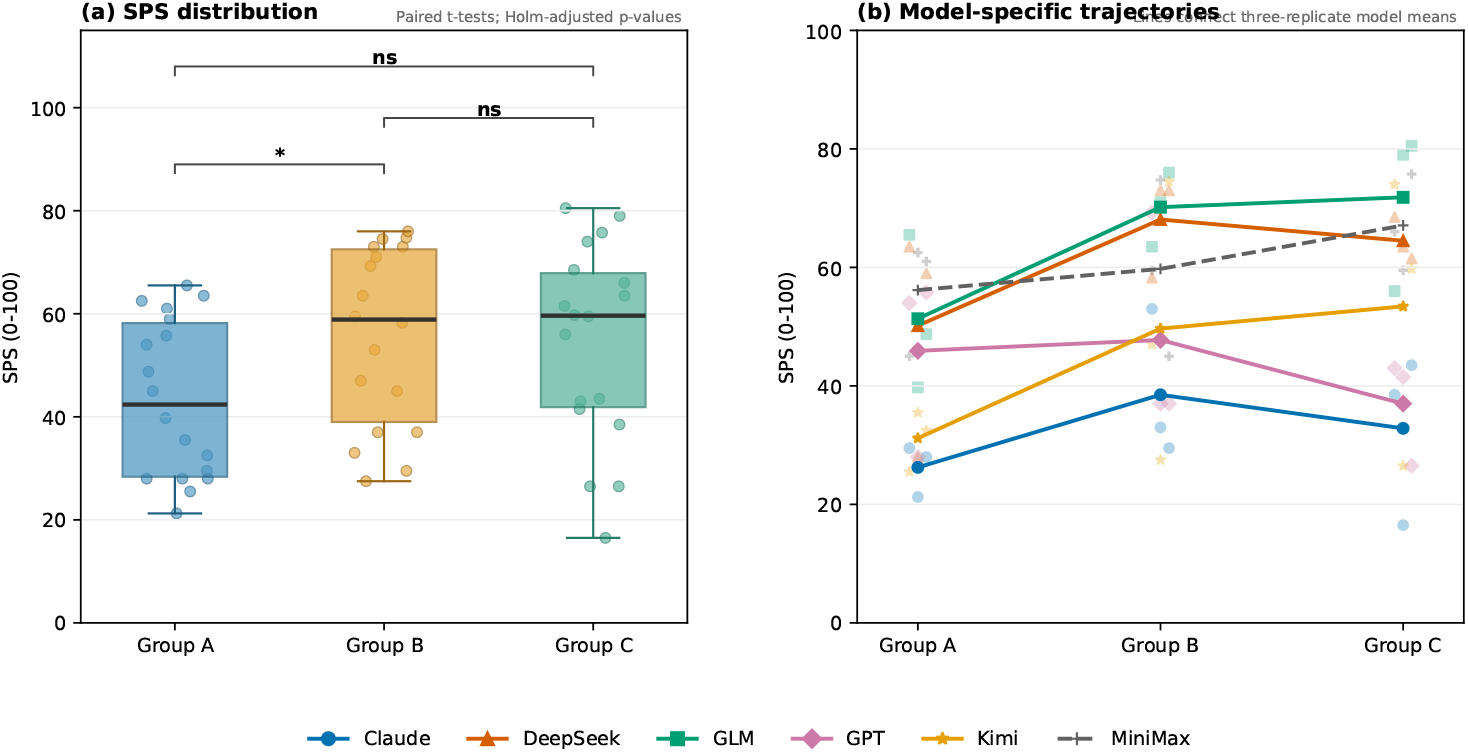
Distribution of SPS across Groups A, B, C and model-specific changes between groups. Points show two-expert mean SPS of individual runs; lines connect each model’s means across Groups A, B, C.

### 3.4 Model performance differs, but no condition-by-model interaction was detected

The two-factor model yielded significant main effects of condition (*P* = 0.023) and of model (*P <* 0.001), but no evidence of a condition-by-model interaction was detected (*F* (10, 36) = 0.603, *P* = 0.801). All six models showed positive *B − A* differences (range +1.83 to +18.83), whereas *C − B* differences were inconsistent in direction (range *−*10.75 to +7.33; Figure 3). The interaction test had limited power given three runs per model per condition.

**Figure 3:**
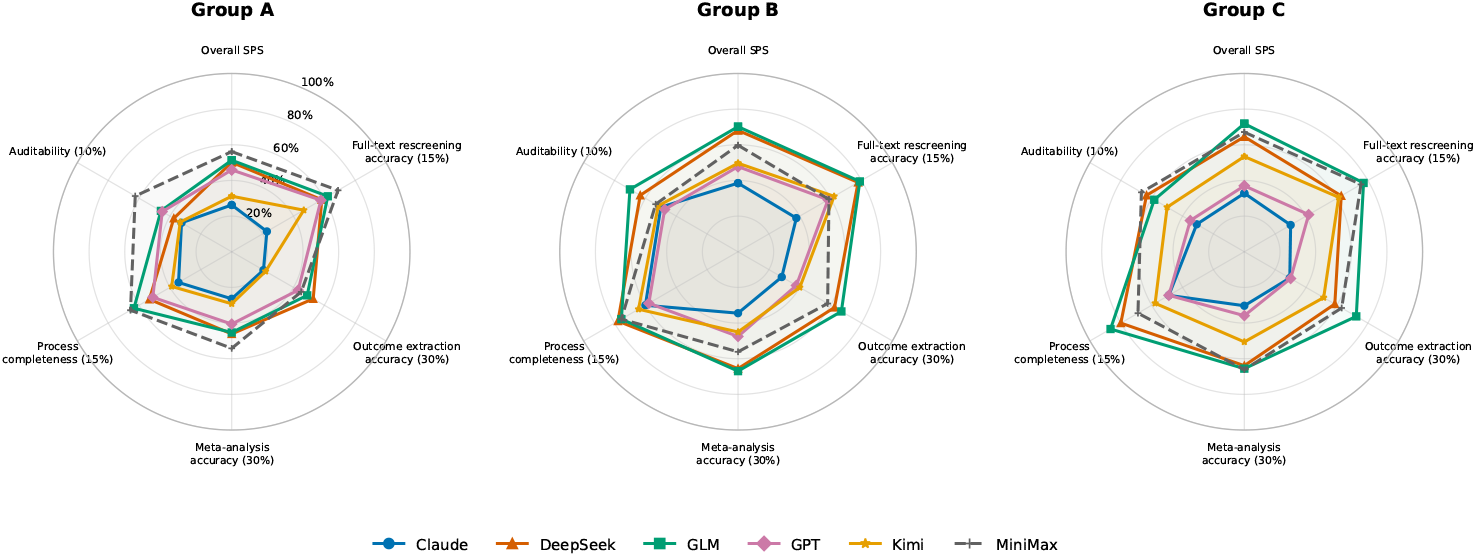
Model-specific SPS profiles across Groups A, B, C. The axes show overall SPS and the five SPS components, each scaled to the percentage of its maximum; values are two-expert means of three runs per model per group. Descriptive only.

### 3.5 SPS component patterns and scientific-risk profiles

All five SPS components increased from condition A to condition B (*B − A*: +1.43 for full-text re-screening and analysis eligibility, +3.07 for the three-outcome data paths, +3.69 for meta-analytic methods and results, +2.61 for core artifacts and analysis completeness, and +1.32 for verifiability and reporting completeness); the component differences do not sum exactly to the overall *B − A* estimate because of rounding. The largest contributions were from the meta-analytic-methods and outcome-data-path dimensions. Across the 54 runs, Expert 2 classified 7 as low, 21 as moderate, 24 as high, and 2 as very high scientific risk; the overall risk-level distribution is reported in this section. No formal component-level inference was performed.

### 3.6 Directional consistency across runs

At the run level, with runs paired by model and replicate number, B scored higher than A in 13/18 pairs and lower in 5/18, and C scored higher than B in 8/18 pairs and lower in 10/18. These counts describe direction only and do not replace effect-size or inferential analysis (Supplementary Figure S3).

### 3.7 Group-specific PFS–SPS associations

Within-condition correlations between PFS and SPS were positive: *r* = 0.680 (95% CI 0.313–0.871; *P* = 0.002) in condition A, 0.856 (95% CI 0.648–0.945; *P <* 0.001) in condition B, and 0.766 (95% CI 0.465–0.908; *P <* 0.001) in condition C. Because PFS-A, PFS-B, and PFS-C are defined on different scales and were scored by the same expert who scored the SPS, these associations are reported as within-condition descriptive results; no pooled slope is reported (Figure 4).

**Figure 4:**
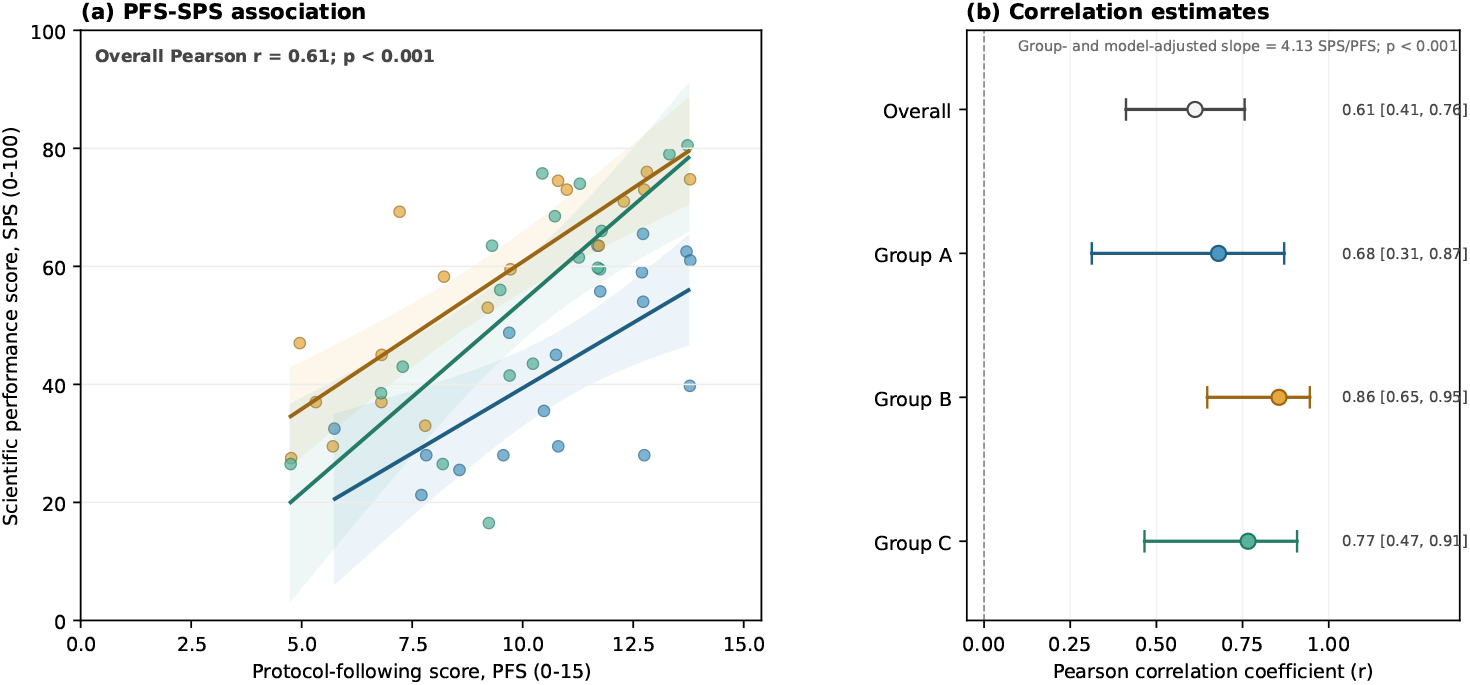
Within-Group relationship between Protocol-following score (PFS) and SPS (two-expert means). Pearson *r* = 0.680 (Group A), 0.856 (Group B), and 0.766 (Group C). An overall *r* = 0.61 with a group- and model-adjusted slope of 4.13 SPS per PFS point is shown for reference; within-condition correlations are the primary report because the PFS scales differ across conditions.

## 4 Discussion

Access to modular skills improved mean scientific performance relative to unassisted execution in this benchmark. Fixed Quality-First orchestration of the same skills did not show an additional overall gain beyond skills, and no condition-by-model interaction was observed. High inter-rater agreement was demonstrated for the scoring framework in this benchmark.

Skills decompose a complex task into explicit, repeatable scientific steps, and structured intermediate products may reduce breaks in the chain linking screening, trial-family identification, extraction, and statistical analysis inputs. The descriptive pattern that *B − A* was positive for all six models supports a broadly consistent direction; however, because the interaction was not significant, we cannot claim that models benefited equally. Our findings align with recent evidence that structured, sub-task-level support can improve LLM-based evidence synthesis [7], and they extend prior skill-based agent evaluations [16] by isolating the incremental effect of fixed orchestration on identical task inputs. Several task-specific features may explain the absence of a detectable incremental benefit. The task pre-specified three formal endpoints, so the prompt itself provided a relatively complete analysis skeleton. Key data were located primarily in body text and tables, which may limit the benefit of enforced image-evidence nodes. Models in condition B could still form their own execution paths, and a fixed DAG may add non-essential steps, increase context consumption, or propagate errors. An alternative explanation is that the current sample and number of repetitions were insufficient to detect a moderate DAG effect: the *C − B* interval still allows a benefit of up to about 6.5 points or a harm of about 8.9 points. We therefore conclude that fixed orchestration provided no detectable incremental benefit in this benchmark, rather than that fixed orchestration is ineffective.

Overall model–configuration performance differed in this benchmark, but the interaction was not significant. Higher baseline capability does not imply larger skill benefits, and lower baseline capability does not imply that a model benefits more from fixed orchestration. Model-specific rises and falls may reflect random variation across three repetitions, context handling, or implementation differences. We do not generalize model rankings to other tasks or future model versions.

The high ICC indicates that the scale can stably distinguish run quality. The observation that Expert 1 scored on average about 2 points higher than Expert 2 suggests residual rater-severity differences, and one expert participated in gold-standard construction and was unmasked; the final scores therefore represent a partially masked, two-expert average.

Within each condition, PFS and SPS were positively correlated, indicating that more complete adherence to the respective task requirements co-occurred with higher SPS. Because the PFS scales differ across conditions, the scores are same-source ratings, and process adherence was not randomized, we do not infer that higher PFS causes higher SPS.

The study compared six models, three conditions, and three independent runs per model per condition on the same task inputs. Conditions B and C used identical skills, separating skill availability from fixed orchestration. Scoring used a study-level gold standard with multiple defensible branches and tolerances rather than language quality alone. All 54 runs were scored by two experts, with explicit label masking for the second expert. Effect sizes, confidence intervals, multiplicity adjustment, and the interaction were reported together.

Several limitations should be considered. First, this benchmark covers a single clinical topic with pre-specified outcomes, and the findings may not generalize to other meta-analysis settings. Second, only three repetitions per model per condition limited the statistical power of model-specific and interaction analyses. Third, the rating design was only partially masked, as Expert 1 was unmasked and constructed the gold standard, while Expert 2 may have inferred experimental conditions from skill artifacts. Future work with more diverse and open-ended tasks, more repetitions, and independent gold-standard experts will help overcome these limitations.

## 5 Conclusion

Across six LLMs completing a controlled meta-analysis benchmark, access to modular skills improved mean scientific performance relative to unassisted execution. Fixed Quality-First orchestration of the same skills did not provide a detectable incremental benefit, and no condition-by-model interaction was observed. High inter-rater agreement was demonstrated for the scoring framework in this benchmark. These findings support modular scientific scaffolding for this task while leaving the value of fixed orchestration unresolved for more open-ended, image-dependent, or heterogeneous evidence-synthesis settings.

## Supporting information

Integrated experimental design and orchestrated meta-analysis workflow showing data flow, dependencies, parallel execution, and conditional diagnostic

Skill data flow across the meta-analysis workflow (Quality-First full path, outcome not predefined).

The 18 workflow nodes (P0, A-Q) of the Quality-First orchestration.

SPS dimensions and maximum points.

## Data Availability

All data produced in the present work are contained in the manuscript

## Declarations

### Author contributions

Huimei Wang and Yingyong Hou conceived and supervised the study and coordinated the project. Yaqing Xiao, Yajun Gu, Pengfei Xia, and Minying Deng contributed equally to this work as co-first authors: Yaqing Xiao organized the meta skills and performed data testing; Yajun Gu developed the meta-automation pipeline; Pengfei Xia led the study design and manuscript arrangement; and Minying Deng performed methodological validation, clinical-content review, and critical revision of the manuscript. Shengyang Xie and Yingzhu Xing contributed equally as co-second authors: Shengyang Xie wrote the manuscript, and Yingzhu Xing conducted the data investigation and contributed to the study design. Haoyue Cheng and Kun Zhu contributed to study-level data checking, review of clinical terminology and outcome definitions, and critical manuscript revision. Qianyu Yao and Fei Sun performed proofreading and manuscript review; Wei Chen and Xueqian Wen reviewed the data; and Yao Chen handled packaging and organization. All authors read and approved the final manuscript.

### Funding

This research received no specific grant from any funding agency in the public, commercial, or not-for-profit sectors.

### Competing interests

The authors declare no competing interests.

### Ethics statement

This study evaluated anonymized AI-generated outputs and publicly available RCT literature; it did not involve patient-level data, clinical samples, or interventions, and no institutional ethics approval was required.

### Data and code availability

Task prompts, skill definitions, the DAG specification, the gold standard, rating rubrics, per-expert scores, and analysis code will be made available upon publication.

### Supplementary material

A single supplementary file accompanies this manuscript and contains the Supplementary Methods (Sections S1–S9), Supplementary Tables S1–S2, and Supplementary Figures S1–S3.

### Generative-AI disclosure

The outputs analyzed in this study were generated by six commercial large language models—GPT-5.5, Claude Sonnet 4.5, GLM-5.2, DeepSeek-V4-Pro, Kimi-K2.6, and MiniMax-M3—accessed through the OpenCode 1.18.9 platform, as described in the Methods. Generative-AI tools were also used to assist with manuscript drafting, literature organization, and figure preparation. All outputs, analyses, references, figures, and manuscript text were independently reviewed and approved by the human authors, who take full responsibility for the final content.

## Supplementary Materials

**Supplementary Figure S1.**
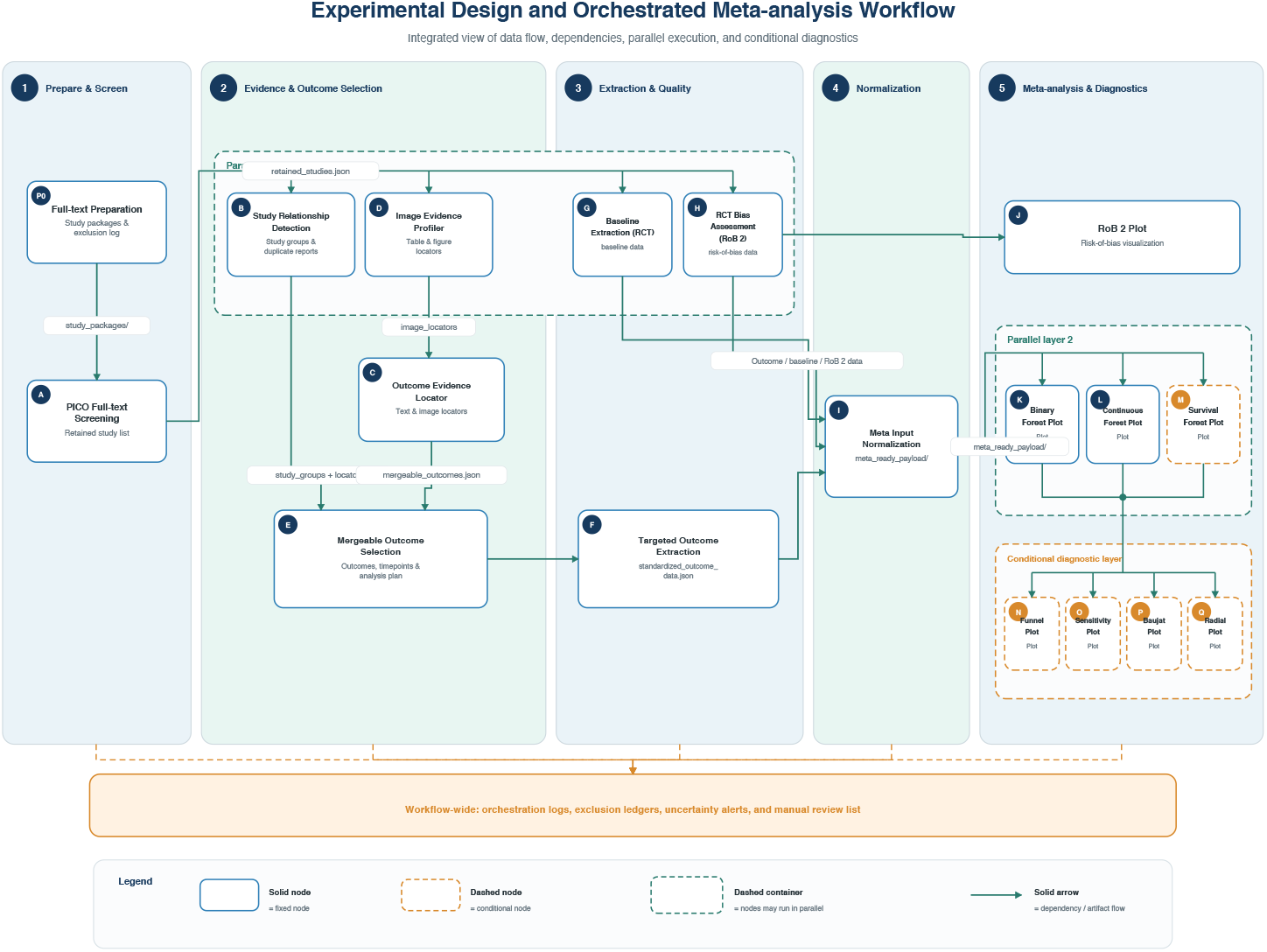
Integrated experimental design and orchestrated meta-analysis workflow showing data flow, dependencies, parallel execution, and conditional diagnostics.

**Supplementary Figure S2.**
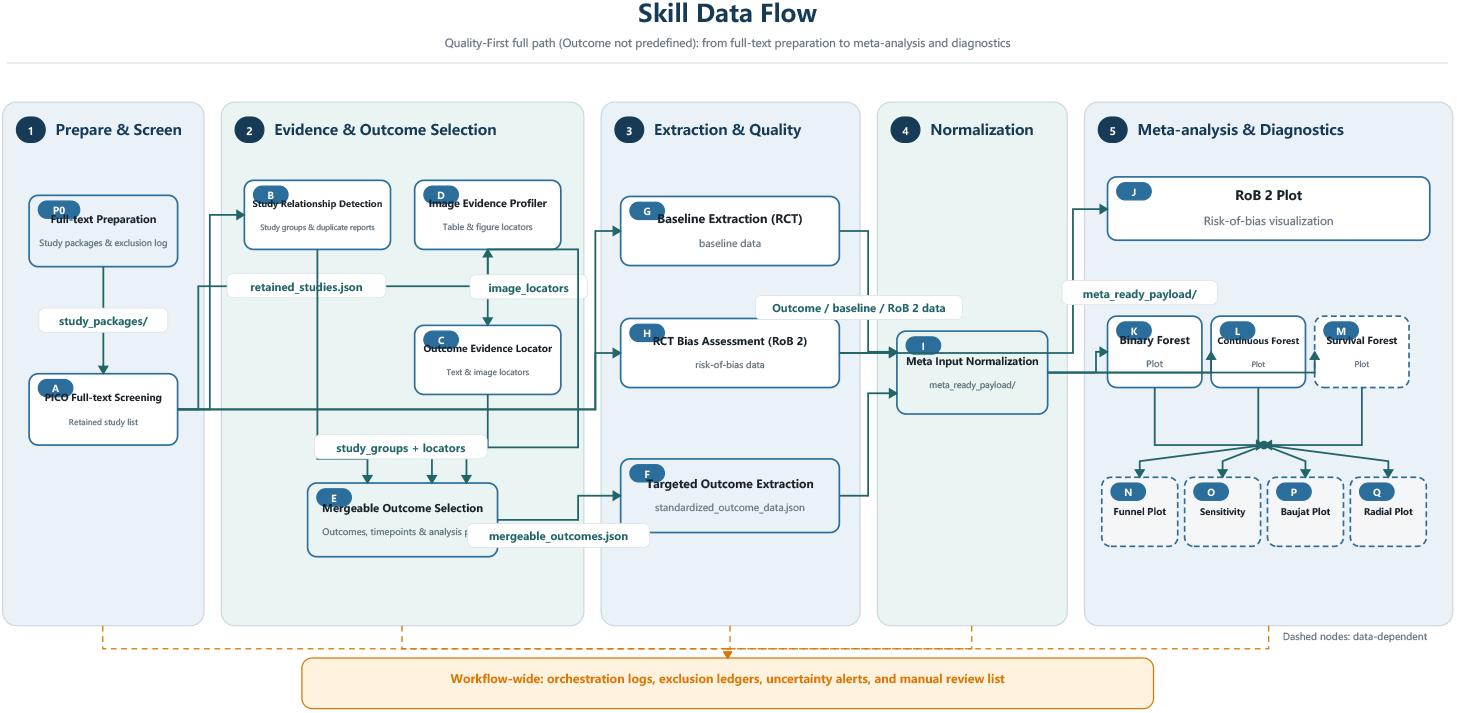
Skill data flow across the meta-analysis workflow.

**Supplementary Figure S3.**
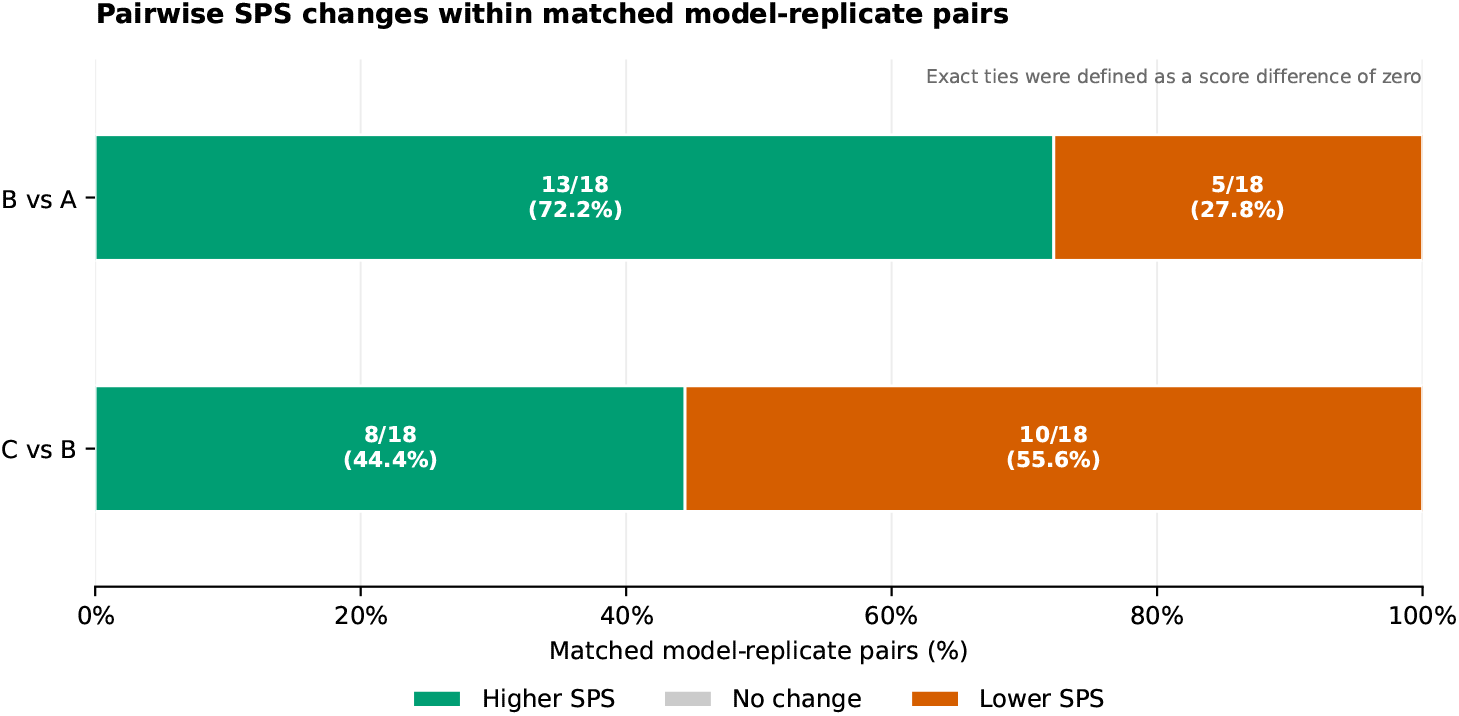
Pairwise win–loss comparison of SPS between conditions across runs, paired by model and replicate number.

## Notes

### Competing Interest Statement

The authors have declared no competing interest.

