## Supplementary material for "Benchmarking modular skills and fixed orchestration across six large language models in a meta-analysis task": Integrated experimental design and orchestrated meta-analysis workflow showing data flow, dependencies, parallel execution, and conditional diagnostic

Integrated view of data flow, dependencies, parallel execution, and conditional diagnostics

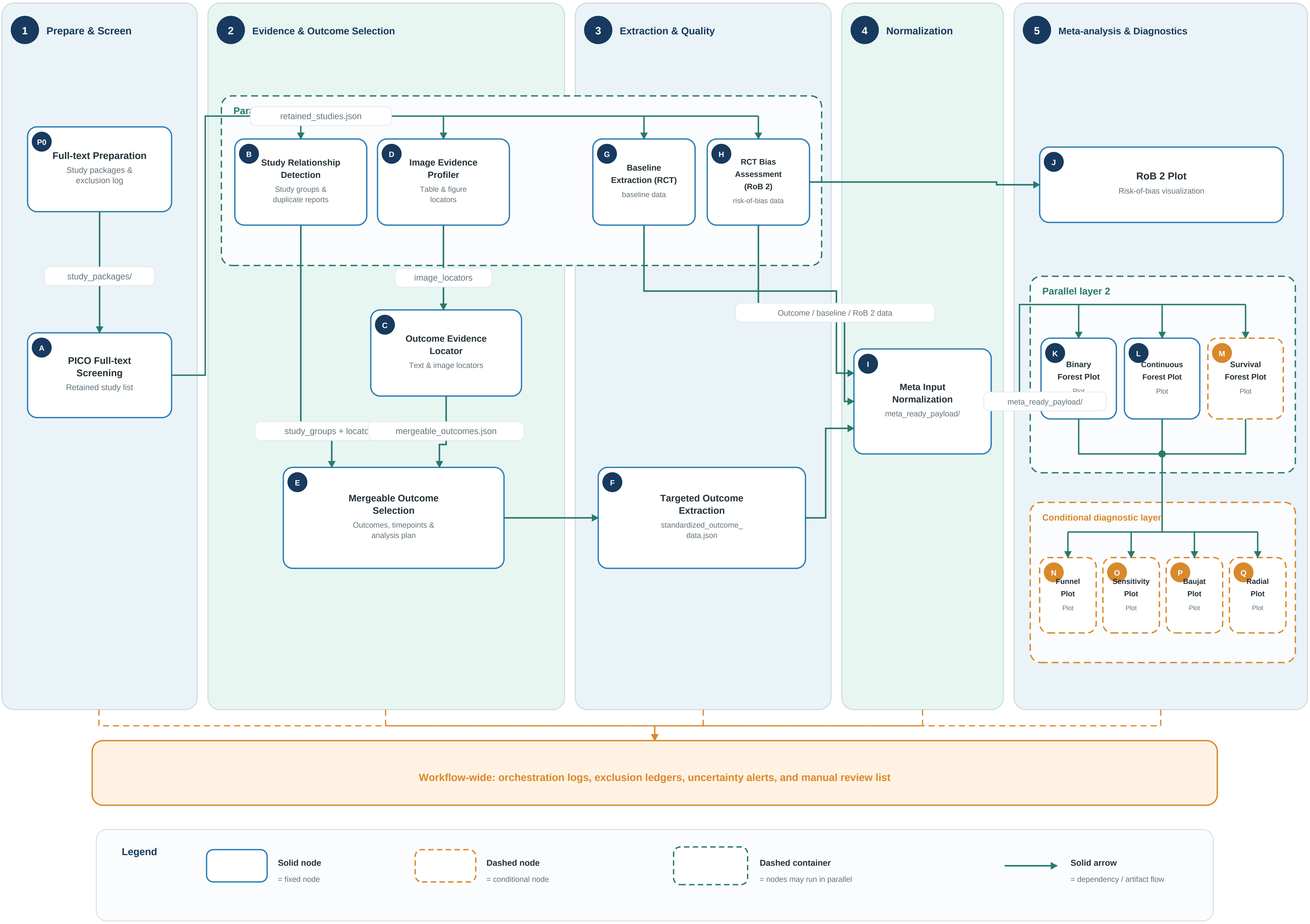
