## Supplementary material for "Benchmarking modular skills and fixed orchestration across six large language models in a meta-analysis task": Skill data flow across the meta-analysis workflow (Quality-First full path, outcome not predefined).

Quality-First full path (Outcome not predefined): from full-text preparation to meta-analysis and diagnostics

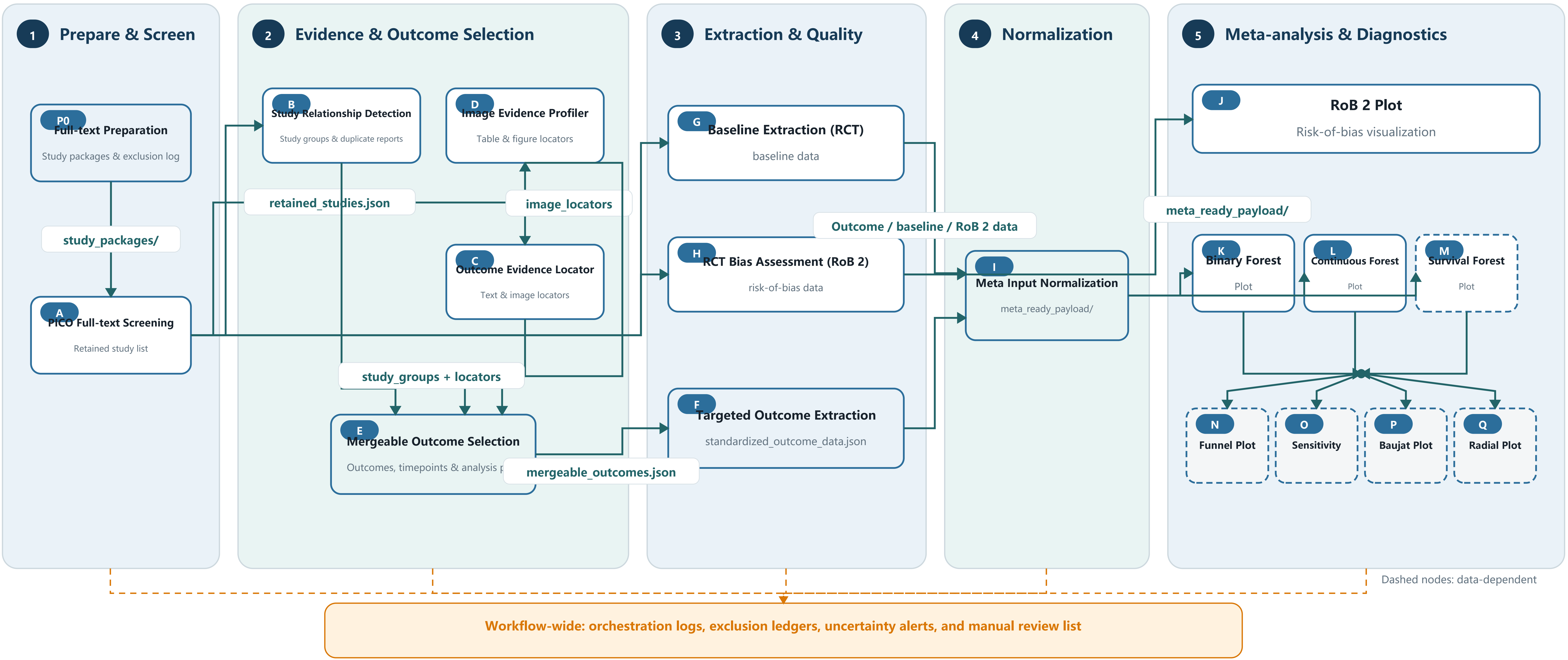
