## Supplementary figures and images for "Benchmarking modular skills and fixed orchestration across six large language models in a meta-analysis task"

### Supplemental Data 1

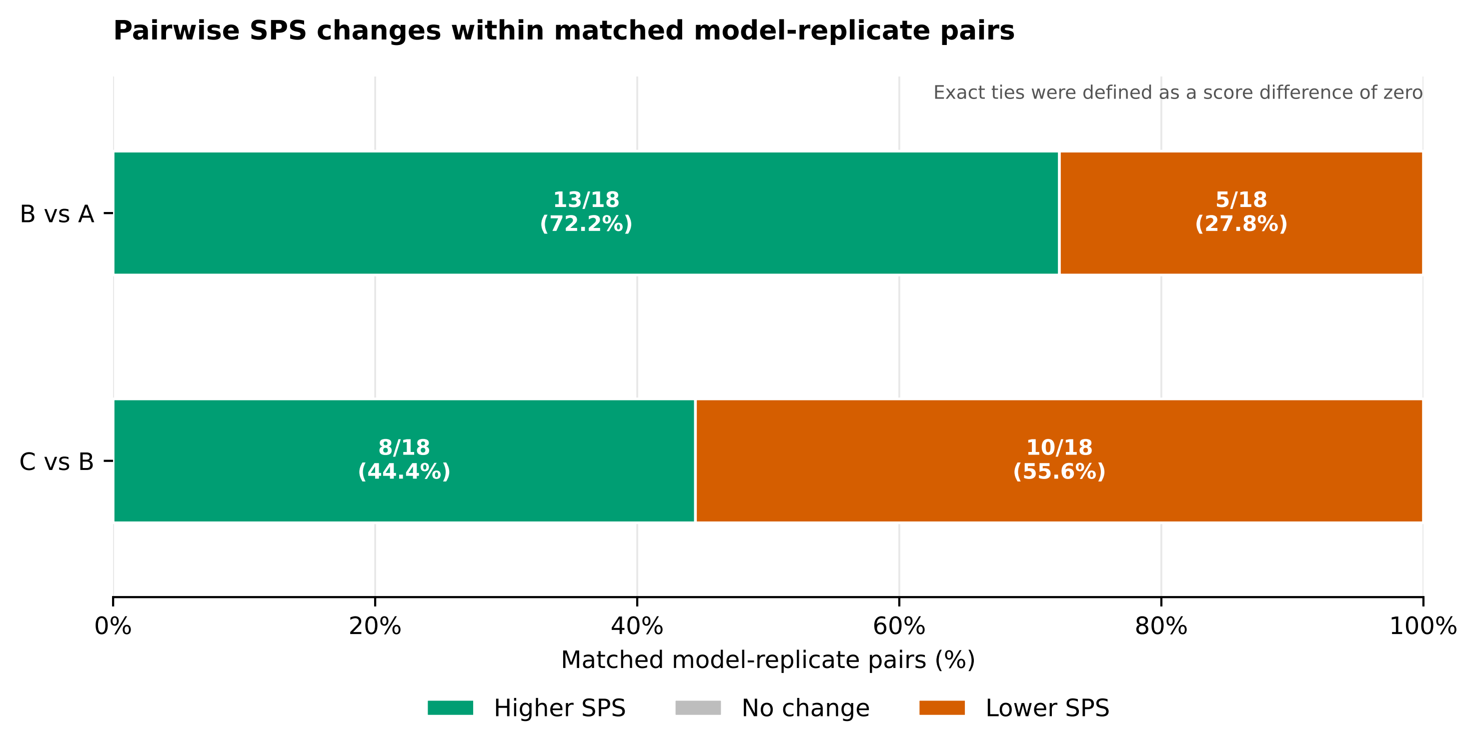
