## Supplementary Materials for "Benchmarking modular skills and fixed orchestration across six large language models in a meta-analysis task"

**Supplementary Methods**

***S1. Modular meta-analysis skill package and node definitions***

The modular skill package covered the functional domains of the meta-analysis workflow: full-text preparation, screening, trial-family identification, outcome location and selection, study-level data extraction, risk-of-bias (RoB 2) assessment, data standardization, meta-analysis, and diagnostic plots. The complete Quality-First workflow comprised 18 nodes (P0 and A–Q) organized in five ordered layers: (1) Prepare & Screen; (2) Evidence & Outcome Selection; (3) Extraction & Quality; (4) Normalization; and (5) Meta-analysis & Diagnostics. Table S1 defines each node, its function within the workflow, and its primary input and output artifacts. Solid nodes denote fixed nodes; dashed nodes denote conditional nodes whose execution depends on the data encountered at run time (e.g., outcome type and data availability); dashed containers denote nodes that may run in parallel.

| **Node** | **Layer** | **Function** | **Primary inputs** | **Primary outputs** |
| --- | --- | --- | --- | --- |
| P0 | 1. Prepare & Screen | Full-text preparation of study packages; exclusion logging | Source RCT reports (20 parsed study packages) | study_packages/; exclusion log |
| A | 1. Prepare & Screen | PICO-based full-text screening | study_packages/ | retained_studies.json (retained study list) |
| B | 2. Evidence & Outcome | Study relationship detection (trial-family identification, duplicate reports) | retained_studies.json | study groups; duplicate-report flags |
| C | 2. Evidence & Outcome | Outcome evidence location in text | retained_studies.json | text locators (image_locators.json) |
| D | 2. Evidence & Outcome | Image evidence profiling (tables and figures) | study groups + locators | table & figure locators |
| E | 2. Evidence & Outcome | Mergeable outcome selection (outcomes, timepoints, analysis plan) | locators | mergeable_outcomes.json |
| F | 3. Extraction & Quality | Targeted outcome extraction | mergeable_outcomes.json | standardized_outcome_data.json |
| G | 3. Extraction & Quality | Baseline extraction (RCT characteristics) | retained_studies.json | baseline data |
| H | 3. Extraction & Quality | RCT risk-of-bias assessment (RoB 2) | retained_studies.json | risk-of-bias data |
| I | 4. Normalization | Meta-analysis input normalization | outcome / baseline / RoB 2 data | meta_ready_payload/ |
| J | 5. Meta-analysis & Diagnostics | RoB 2 visualization | meta_ready_payload/ | risk-of-bias plot |
| K | 5. Meta-analysis & Diagnostics | Binary-outcome forest plot | meta_ready_payload/ | forest plot (binary) |
| L | 5. Meta-analysis & Diagnostics | Continuous-outcome forest plot | meta_ready_payload/ | forest plot (continuous) |
| M | 5. Meta-analysis & Diagnostics | Survival-outcome forest plot | meta_ready_payload/ | forest plot (time-to-event) |
| N | 5. Meta-analysis & Diagnostics | Funnel plot | meta_ready_payload/ | funnel plot |
| O | 5. Meta-analysis & Diagnostics | Sensitivity analysis plot | meta_ready_payload/ | sensitivity plot |
| P | 5. Meta-analysis & Diagnostics | Baujat plot | meta_ready_payload/ | Baujat plot |
| Q | 5. Meta-analysis & Diagnostics | Radial plot | meta_ready_payload/ | radial plot |

*Table S1. The 18 workflow nodes (P0, A–Q) of the Quality-First orchestration.*

***S2. The Quality-First fixed directed acyclic graph (DAG)***

The fixed orchestration defined (i) parallel layers, in which independent nodes (e.g., B, C, and D in layer 2) may execute concurrently; (ii) serial dependencies between layers, such that downstream nodes can start only after their upstream inputs become available; and (iii) conditional nodes, which execute only when the required data conditions are met (for example, binary, continuous, or survival forest plots are generated according to the outcome type actually present in the extracted data). Workflow-wide, the orchestrator maintained orchestration logs, exclusion ledgers, uncertainty alerts, and a manual-review list. The complete DAG is shown in Supplementary Figure S1.

***S3. Skill data flow***

Supplementary Figure S2 shows the skill data flow along the Quality-First full path when the target outcome is not predefined: full-text preparation (P0) and screening (A) are followed by study-relationship detection (B), outcome-evidence location (C), and image-evidence profiling (D); mergeable outcome selection (E) then feeds targeted outcome extraction (F), alongside baseline extraction (G) and RoB 2 assessment (H); normalized meta-analysis inputs (I) feed the diagnostic layer (J–Q). Each artifact is passed between nodes as a structured file, so that the chain from screening to pooled estimate remains traceable.

***S4. Benchmark task specification***

The benchmark task was a pre-specified randomized controlled trial (RCT) meta-analysis comparing robotic-assisted surgery with conventional laparoscopic surgery for colorectal cancer (population: patients with colorectal cancer; intervention: robotic-assisted surgery; comparator: conventional laparoscopic surgery; eligible designs: randomized controlled trials). The input comprised 20 fully parsed study packages derived from the full texts of published RCT reports. Three outcomes served as formal endpoints for scoring: conversion to open surgery, operative time, and number of lymph nodes harvested. Postoperative complications were collected but evaluated only descriptively and did not contribute to SPS scoring, score caps, or scientific-safety classification.

***S5. Gold standard construction and defensible branches***

A study-level gold standard was constructed by Expert 1 (a long-term meta-analysis researcher) and independently reviewed within the study team. The gold standard allowed multiple defensible analysis branches rather than a single correct path. Specifically, three branch families were pre-specified: (i) a language-policy branch, permitting inclusion or exclusion of non-Chinese/non-English studies according to a pre-specified language policy; (ii) a continuous-data branch, permitting inclusion of continuous data with or without transformation; and (iii) a trial-family branch, requiring mutually exclusive handling of REAL/Tang criteria reports versus parent-study/sub-study reports. For each acceptable branch, the corresponding study set, meta-analysis inputs, pooled effect, and tolerance ranges were predefined, and extraction was scored against study-level reference data with these tolerances.

**Per-branch study lists, reference values, pooled effects, and tolerance ranges for the three formal endpoints Detailed per-branch study sets, reference values, pooled effects, and tolerance ranges are available from the corresponding author upon reasonable request.**

***S6. Scientific Performance Score (SPS)***

The SPS comprised five weighted dimensions summing to 100 points (Table S2). All 54 runs were scored by two meta-analysis experts; Expert 2 was masked to model and condition labels during SPS scoring, and final scores were the arithmetic mean of the two raters.

| **Dimension** | **Maximum points** | **Scope** |
| --- | --- | --- |
| Full-text re-screening and analysis eligibility | 15 | Correct re-screening of the 20 study packages and application of eligibility criteria under the chosen defensible branch |
| Study-level data and analysis paths for the three formal outcomes | 30 | Correct study-level extraction and analysis inputs for conversion to open surgery, operative time, and lymph nodes harvested |
| Meta-analytic methods and results | 30 | Appropriate pooling methods (effect measure, model, heterogeneity assessment) and correctly computed pooled estimates within tolerance |
| Core artifacts and analysis completeness | 15 | Generation of the required analysis artifacts (data tables, forest plots, diagnostics) and completeness of the analysis chain |
| Verifiability and reporting completeness | 10 | Traceability of data paths and completeness of reporting sufficient for independent verification |
| Total | 100 | — |

*Table S2. SPS dimensions and maximum points.*

**Item-level scoring rubric, score-cap rules, and worked examples The item-level rubric and any score-cap rules applied during scoring are available from the corresponding author upon reasonable request.**

***S7. Protocol-Following Score (PFS)***

Group-specific Protocol-following scores (PFS-A, PFS-B, PFS-C), each scored out of 15 points, measured how completely each run followed the process requirements of its own condition. PFS-A assessed compliance with the requirement not to use meta-analysis-specific skills, task boundaries, and method instructions. PFS-B assessed the actual invocation, outputs, and upstream/downstream transfer of individual skills. PFS-C assessed adherence to the Quality-First orchestrator, DAG dependencies, and data transfer at key nodes. The three scores are condition-specific, are not directly comparable across conditions, and are not summed with the SPS.

***S8. Scientific safety/risk classification***

Both experts independently assigned each run a scientific safety/risk level (low, moderate, high, or very high) according to explicit criteria based on the scientific-safety bottom line. Risk levels reflected the scientific consequences of acting on the run's conclusions (for example, materially wrong pooled estimates, untraceable data paths, or unfounded conclusions). Expert 2 classified the 54 runs as 7 low, 21 moderate, 24 high, and 2 very high; agreement statistics are reported in the main manuscript.

**Full operational criteria for the four risk levels The complete risk-level criteria wording used by the two raters are available from the corresponding author upon reasonable request.**

***S9. Masking, de-identification, and rating batches***

All 54 runs were scored independently by two experts. Expert 1 was aware of models and conditions and participated in gold-standard construction; Expert 2 was masked during SPS scoring. Runs were presented to Expert 2 under random identifiers B001–B054. Explicit model and condition labels were removed from run texts and from text embedded in PNG files; time metadata and file timestamps were also removed. The identifier mapping was generated from a confidential seed and path-based SHA-256 ordering, and runs were delivered in nine masked rating batches. Source and masked directories were cross-checked by SHA-256, with de-identification logs retained. PFS scoring was not masked because process evidence (skills and DAG logs) had to be visible to the rater.

**Supplementary Figures**

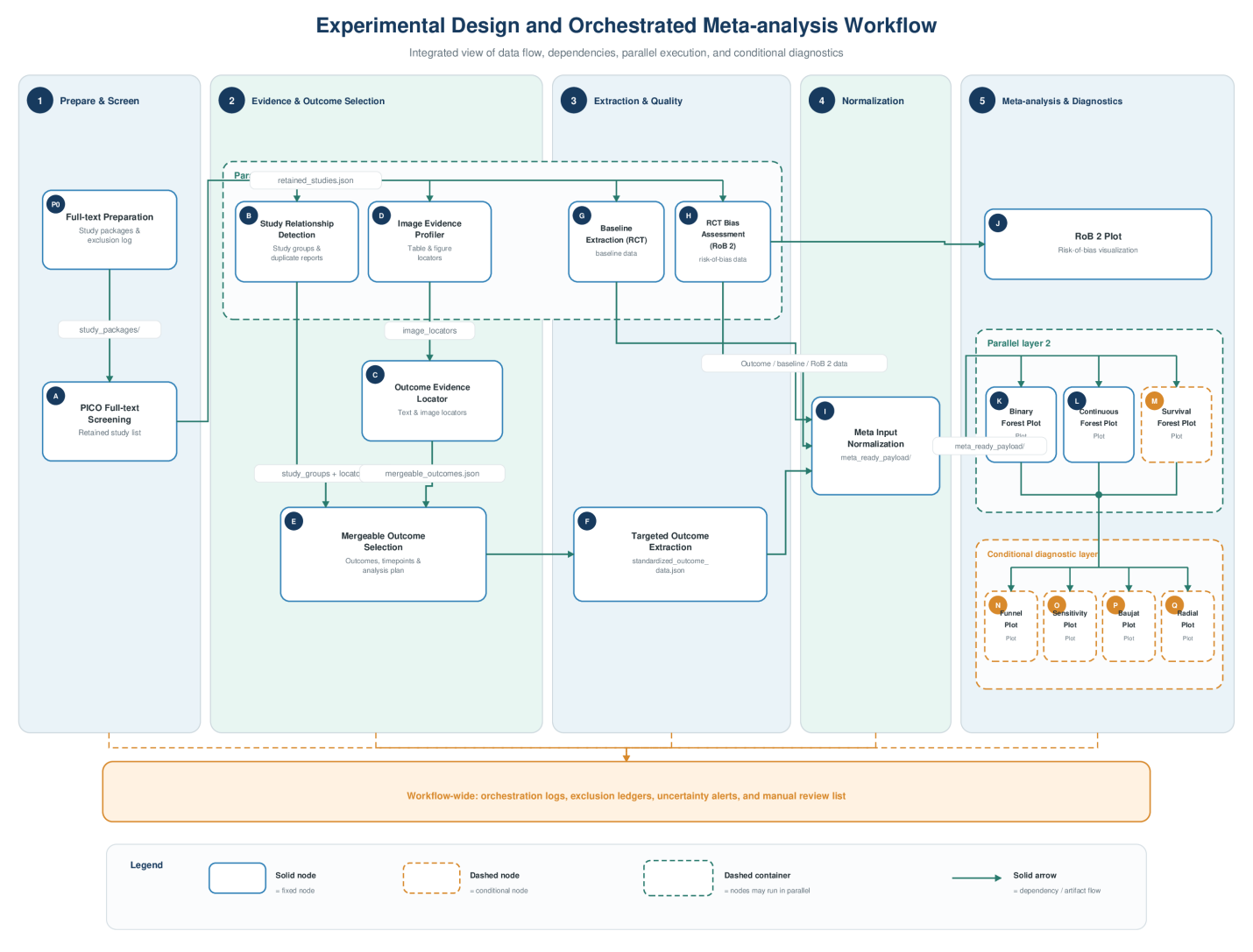

*Supplementary Figure S1. Integrated experimental design and orchestrated meta-analysis workflow showing data flow, dependencies, parallel execution, and conditional diagnostics.*

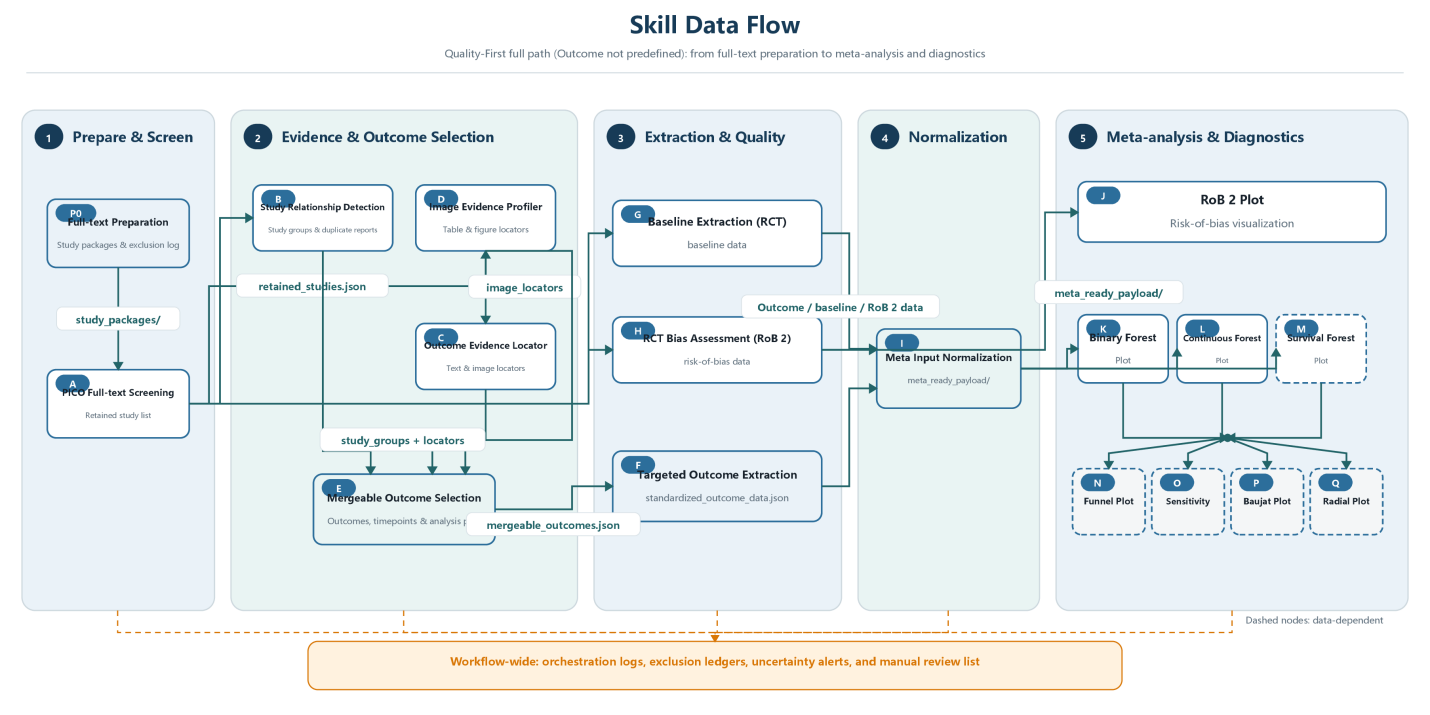

*Supplementary Figure S2. Skill data flow across the meta-analysis workflow (Quality-First full path, outcome not predefined).*

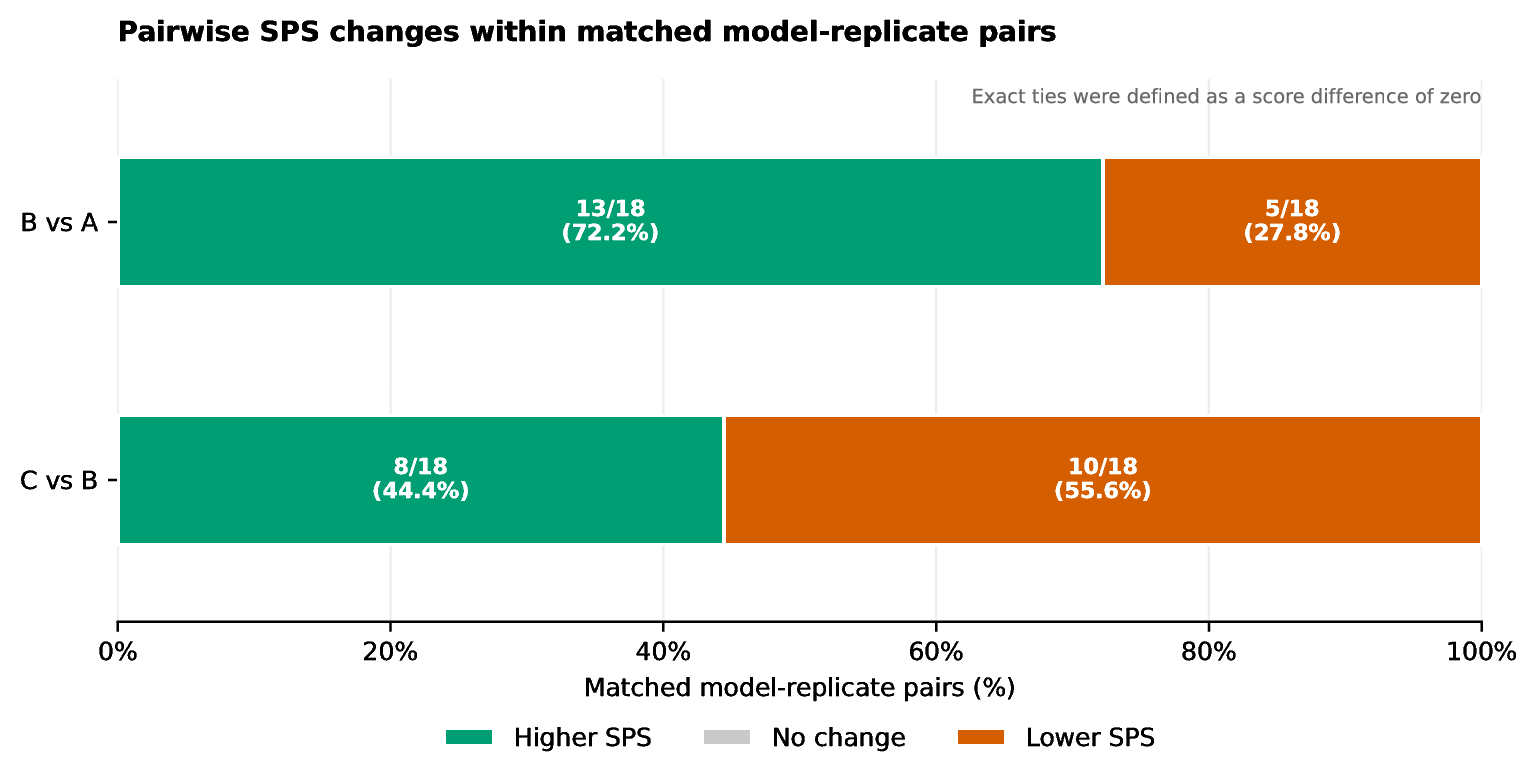

*Supplementary Figure S3. Pairwise win–loss comparison of SPS between conditions across runs, paired by model and replicate number.*
